# What is the effectiveness of antiracist interventions for ethnic minority healthcare staff? A systematic review

**DOI:** 10.1101/2024.06.13.24308817

**Authors:** Kismet Lalli, Oluwatomisin Adewole, Holly Peters, Chris Price, Blanche Lumb, Micaela Gal, Alison Cooper

## Abstract

**Background:** The National Health Service (NHS) has the most diverse workforce in the United Kingdom (UK), 25% (n= 309,532/1,200,000) of staff belong to ethnic minority groups. However, there is evidence of longstanding issues of racism within the NHS and discrimination towards ethnic minority healthcare staff has been rising since 2016. In the first wave of the COVID-19 pandemic, 95% of COVID-19 deaths among doctors were in an ethnic minority group. There has been no definitive answer for the disproportionate COVID-19 mortality but socioeconomic factors due to structural racism have been suggested as the main drivers. No studies have assessed the effectiveness of antiracist interventions for healthcare staff.

**Methods:** We conducted a systematic review; databases searched included: AMED, Medline via OVID, CINAHL, APA Pyscinfo, Web of Science and OVID Emcare 25^th^– 31^st^ January 2022. The interventions were structured using a model of antiracist interventions and analysed using narrative synthesis methods.

**Results:** 16 papers were reviewed with interventions at different levels: personally mediated (n=9), multilevel (n=4) and institutional (n=3). Personally mediated interventions were workshops (n=8) and a mentorship scheme (n=1). Institutional interventions were policies (n=2) and increasing diversity initiative (n=1). Multilevel interventions were a mix of both. Study designs and risk of bias tools indicated that the quality of evidence was of low quality. Only two studies included control groups. Countries included the USA (n=11), Canada (n=1) and the UK (n=4).

**Conclusion:** There is a lack of robust evidence for antiracist interventions for healthcare staff, especially at an institutional level. High quality research is required to evaluate the long-term effects of interventions.

**Funding statement:** The Wales COVID-19 Evidence Centre was funded for this work by Health and Care Research Wales on behalf of Welsh Government.

## Introduction

The National Health Service (NHS) has the most ethnically diverse workforce in the United Kingdom (UK), with 21% of staff belonging to ethnic minority groups (1). However, racism towards staff operates at many levels; from bullying and harassment (2), to limited career advancement (3), and increased disciplinary action (4). During the first COVID-19 wave (year), 95% of all doctors who died were from ethnic minorities (5). There has been no definitive answer for this disproportionate COVID-19 mortality. Socio-economic factors, such as deprivation, due to longstanding structural racism were cited as main contributors to this mortality rate and the effectiveness of interventions addressing this is unclear (6).

Institutional racism can be defined as ‘the collective failure of an organisation to provide an appropriate service to people based on their ethnic origin, evident in processes, attitudes and behaviours amounting to discrimination, disadvantaging people in ethnic minority groups’ (7). In 1999, the NHS established ‘The National Plan for Action to Tackle Racial Harassment (8)’, and diversity training and equal opportunities (EO) policies have existed for the past 30 years (9).

However, NHS Workforce Race Equality Standard (WRES) data show Black Asian and Minority Ethnic (BAME) staff experiences of harassment and discrimination from colleagues and patients has been increasing since 2016 (2, 10).

An antiracist outlook accepts that racial inequities are rooted in power and policy and works to dismantle systems which have racism embedded within (11). Therefore, an antiracist intervention tackles multiple levels within the system and is “action oriented” (12). We identified a paper reviewing antiracist interventions in outpatient healthcare settings which presented a conceptual model for implementing anti-racism interventions in healthcare at the personal or institutional level (see Figure 1) (13). Publications in recent years that focus on antiracist interventions in a healthcare setting include opinion pieces (14-19), literature reviews (9, 20-24) or studies focussed on patients (13, 25-27). Currently, there is no systematic review assessing the effectiveness of antiracist interventions for healthcare staff.

**Figure 1.**
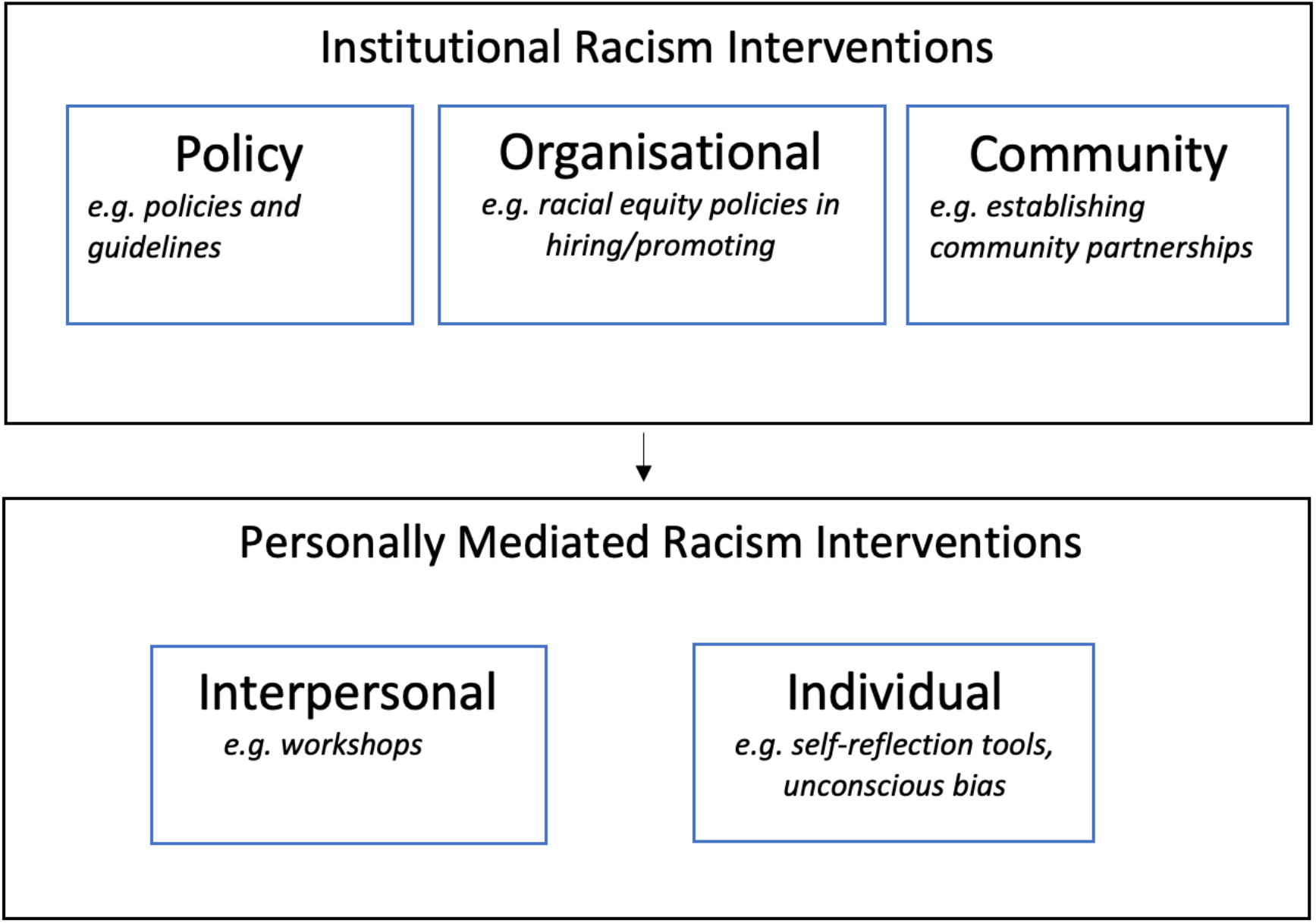
Adapted conceptual model for implementing anti-racist interventions in healthcare (13).

Our aim was to conduct a systematic review and describe the effectiveness of antiracist interventions for healthcare staff. Our objectives included:

1. Identifying interventions addressing racism in healthcare systems in ‘Organization for Economic Cooperation Development’ (OECD) countries
2. Classifying the interventions level based on our adapted conceptual model for implementing anti-racist interventions in healthcare (Figure 1)
3. Evaluating the effectiveness of these interventions; and

## Methods

This review was registered with the International Prospective Register of Systematic Reviews (PROSPERO) and conducted in accordance with the Preferred Reporting Items for Systematic Reviews and Meta-Analysis (PRISMA) guidelines (28), following the framework outlined in the Cochrane Handbook for Systematic Reviews of Interventions (29). Stakeholders from the General Medical Council (KL, BL) and Health Education Improvement Wales (CP, RJ) supported the project and assisted with clarification of terminology, protocol planning, review progress meetings and dissemination of findings. Four meeting were held from July 2021 to April 2022.

### Eligibility criteria

We developed the eligibility criteria (Table 2) in consultation with the stakeholders and the supervisors. As this is the first review focussed on antiracist interventions for healthcare staff, the population, outcome and study design criteria were purposefully broad to map all available evidence. Only evaluated interventions were included as the research question was focused on effectiveness.

**Table 2.**
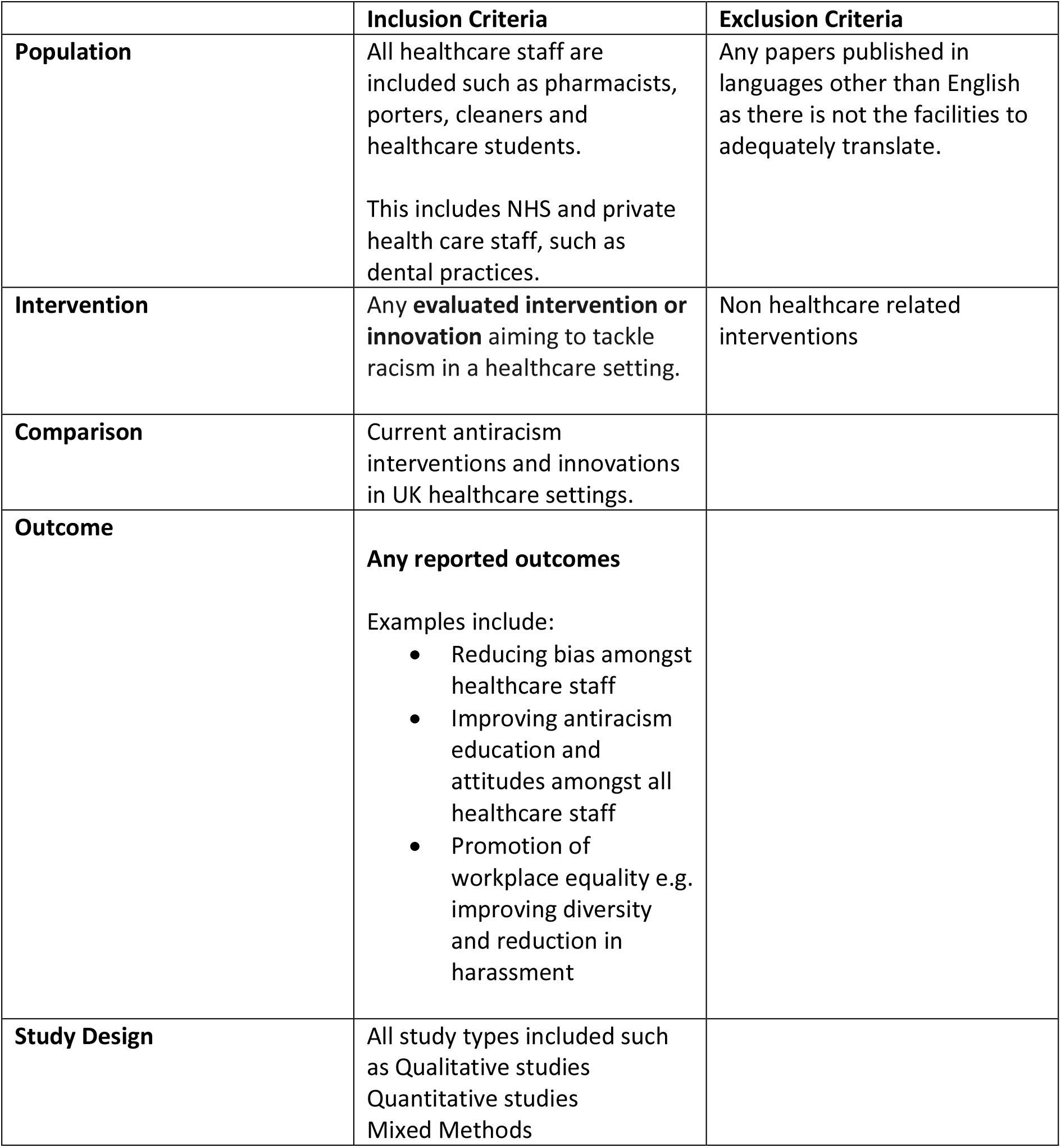
Study eligibility criteria.

### Search Strategy

We developed the search strategy in consultation with information scientists (DM, EG), project supervisors (AC, MG, AE and HP) and stakeholders (KL, BL, CP, RJ). Search terms used key concepts from the research question: ‘racism’, ‘healthcare staff’ and ‘ethnic minority groups’ (found in supplementary material). No restrictions were placed on publication date and non-English language studies were excluded.

### Data sources

Databases searched were Medline via OVID, Ovid Emcare, PsycINFO, CINAHL, Web of science, AMED from 25^th^– 31^st^ January 2022.

### Selection process

Both title and abstract and full text screening had 20% dual screened by another reviewer (TA) as per the Cochrane rapid review guidelines (30). Any uncertainty about paper inclusion was discussed with the co-authors (AC, MG, HP). Aside from searching the databases, citation searching of relevant papers was undertaken. The final included papers were exported to Microsoft Excel spreadsheets for data extraction.

### Data analysis

The included studies were mapped onto the conceptual model to categorise whether they targeted the institutional or personal racism level.

### Critical appraisal

Most of the studies did not use robust analytic study designs, hindering the selection of an appropriate critical appraisal checklist. Study descriptors were assigned, based on the closest match, using an algorithm for assessing appropriate research design as the authors’ descriptors were unclear (31).

Eight studies were appraised using a quality assessment tool for before-after (pre-post) studies with no control group developed by the National Heart, Lungs and Blood Institute (NHLBI) (32). These study designs were unclear and the algorithm identified them as repeat cross-sectional pre-post non-intervention, making the NHLBI checklist the most appropriate. One further study was appraised using the NHLBI non experimental cross-sectional checklist (32). Three qualitative papers were appraised using the Joanna Briggs Institute (JBI) qualitative checklist (33) and one using the JBI checklist for quasi-experimental studies (34). The mixed methods appraisal tool (MMAT) was used for three mixed-methods studies (35).

## Results

Six databases identified 2122 records with 600 duplicates removed. The remaining 1522 records were title and abstract screened, of these 103 were full text screened for eligibility. An additional three papers were identified from citation tracking (see Figure 2). A total of 16 studies was included: qualitative (n=3), quasi-experimental (n=1), mixed methods (n=3) and observational (n=9). Countries included the USA (n=11), Canada (n=1) and the UK (n=4). Most evaluated interventions were aimed at the personally mediated level (n=9), with some multilevel (n=4) and institutional (n=3), based on the adapted model.

**Figure 2.**
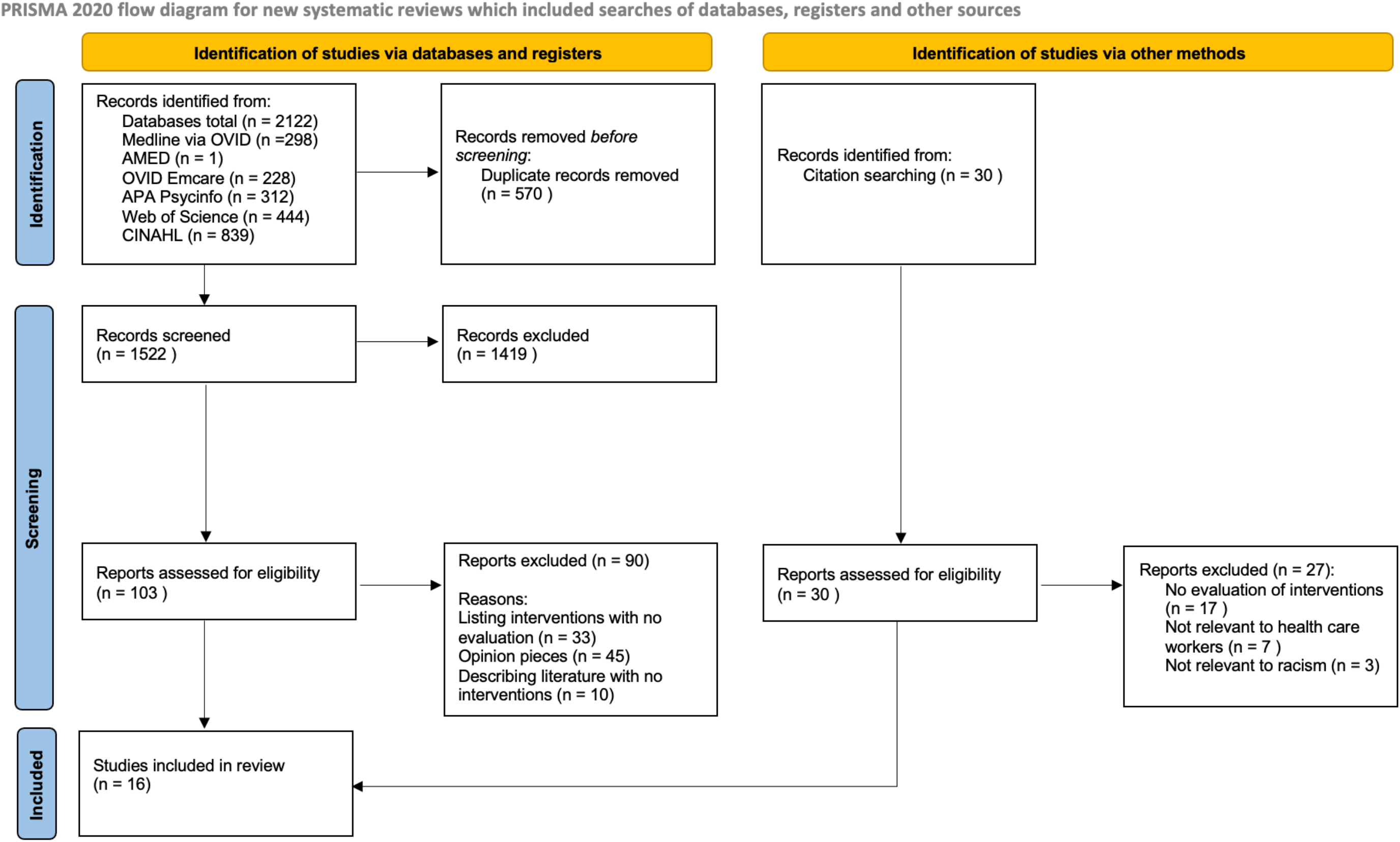
PRISMA Flowchart

Study designs and risk of bias tools indicated that the quality of evidence was of low quality. Only two studies included control groups (36, 37). Most studies could not confidently attribute positive outcomes to the intervention and the results had limited transferability due to small sample sizes. Both qualitative papers examining the success of policy implementation had high risks of bias; the methodology was unclear and neither study acknowledged the researcher’s role. Figure 3 highlights the lack of evaluated institutional antiracist interventions with no interventions identified at a community level.

**Figure 3.**
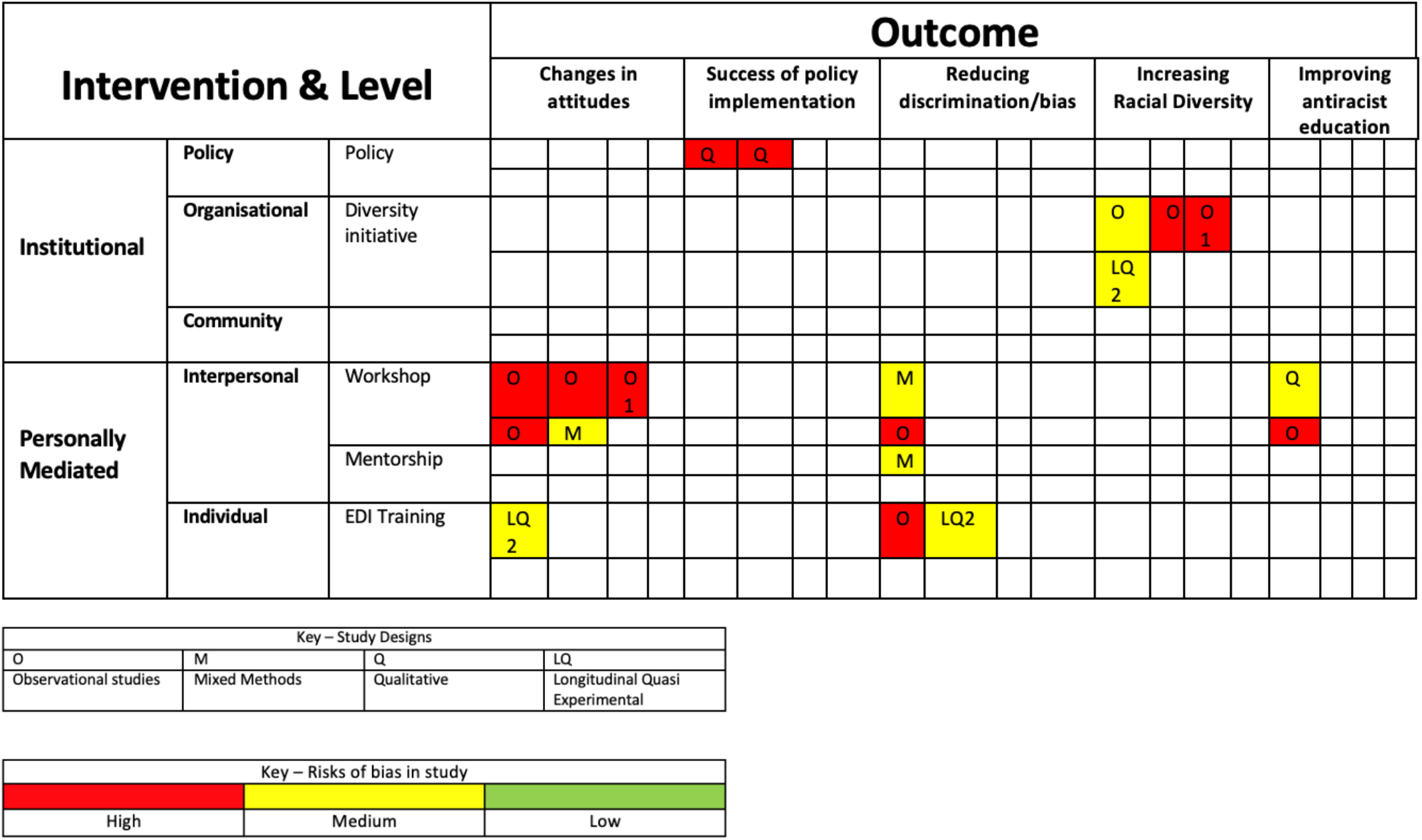
Evidence Map of intervention type, study design and outcomes

### Institutional Interventions

One paper assessing increasing diversity initiatives (38) and two papers evaluating the effectiveness of policies regarding equal opportunities and ethnic diversity respectively (39, 40) were reviewed.

#### Diversity Initiatives

In 2019, Wusu et al published an American observational study aiming to increase underrepresented minority (URM) physicians in a residency programme (38). The interventions included: outreach to URM candidates, revising interviews to minimise bias and ongoing analysis of recruitment data. The study ran from 2014-2017 and URM applicants increased by 80%, a statistically significant increase (P<0.001). However, there was no control group and limited generalisability as it was one family medicine programme in Boston. The positive results cannot be solely attributed to the intervention as this study design was low quality with high risk of bias.

#### Policy Evaluation

There were two papers that evaluated policy. However, both were over 20 years old and did not follow formal qualitative research methodology, therefore these were at risk of bias.

The evaluation of the equal opportunity policy was conducted by interviews with 50 ethnic minority staff and 22 managers at one NHS institution in 1999 (39). The interviews demonstrated the managers lacked knowledge about EO policy and procedures. Despite them all being responsible, only six could identify an EO policy. None received EO training before their role which was attributed to ‘the urgent driving out the important,’ as there were other priorities. Staff interviews highlighted the meaning of EO policies differed between the groups; staff understanding of EO policies centred on career development and promotion, whereas managers focused on recruitment. The paper suggested that these contrasting views may contribute to the disproportionate presence of ethnic minorities in lower grade jobs.

In 2004, the perceptions of ethnic diversity policy in the NHS were evaluated through a mixed methods qualitative and quantitative study (40). A mail survey of every NHS trust was sent out with a response rate of 58% and 32 semi-structured interviews were conducted at a hospital in the south-west of England with employees in charge of employment decisions. Greater diversity was linked to enhanced working relationships and staff perceived an improved service quality. Staff recognised that ethnic minority groups are confined to the lowest paid jobs and that the ultimate goal of ethnic diversity policy should be diversification of senior NHS policymakers as they ‘set the culture of the organisation.’ Staff were supportive of the principles of ethnic diversity policy but unsure how it translates to practice.

#### Personally mediated Interventions

Of the nine personally mediated interventions, eight were workshops and one was a mentorship scheme. Three workshops had supplementary adjuncts, such as drama teaching.

#### Singular Workshops

Six discussion-based workshops followed similar structures of a lecture about the topic then group work. One was UK-based (41), one from Canada (42) and four from the US (43-46). The participant numbers varied and were based on opportunistic sampling.

Workshop topics included: racial harassment, microaggressions, cultural competency, equity and inclusion and implicit bias. They all reported positive responses from post intervention surveys. Participants reported improved knowledge and understanding of anti-racist strategies (42, 45). Four papers highlighted knowledge gaps that require ongoing antiracist education – not single workshops (41, 43, 46, 47). They all employed basic study designs with no control group or long-term follow-up. Bheenuck et al recognised this and recommended future research should assess intervention sustainability (41).

Two studies conducted in 2010 and 2021 respectively reported that the interventions were not entirely successful. In Steed’s 2010 mixed methods qualitative and quantitative study, 13 white female occupational therapists, based in Louisiana, participated in a six-hour cultural competency workshop which aimed to reduce racial bias (46). Their attitudes were measured pre- and post-intervention using the racial arguments scale (RAS) and racial attitudes implicit association test (RAIAT). The workshop produced no significant change in racial attitudes. It was noted that the group held strong negative attitudes towards African Americans in the pre-intervention tests (46). These results suggest individuals with negative views require a different approach and may benefit from longer duration projects. However, the wider generalisability is limited due to the small, homogenous sample.

In 2020, an observational study testing the effect of active learning workshops on reducing implicit bias was evaluated on 137 first year American medical students (44). The Implicit Association Test (IAT) was used pre- and post-intervention to measure the students’ bias. The workshop success differed among racial groups. White students showed positive pre- and post-workshop changes yet Southeast Asian and Asian American students did not show a significant reduction in implicit stereotyping (44). The results suggests that antiracism interventions may need to be adapted for individuals’ backgrounds. It is important to note that the IAT is not a validated tool for measuring implicit bias and for first time users, the reliability is much lower due to unfamiliarity (48), therefore this may have affected the results.

#### Workshops with Adjuncts

Supplemented workshops included a virtual reality experience (49), an interactive map charting locations representing systemic racism in the US (47) and dramaturgical teaching (43). The virtual reality session reported positive outcomes with 94.7% of participants agreeing it was a useful tool for enhancing empathy. The interactive map of racism was acknowledged as a ‘powerful tool’ that improves awareness and shows systemic racism is undeniable. Re-enactment of racial harassment and discussion of how to handle these situations led to positive responses, 100% of participants reported increased confidence in initiating conversations about race.

#### Mentorship scheme

Naidoo’s 2021 mixed methods qualitative and quantitative study piloted a mentorship scheme for eight ethnic minority physical therapy students in the USA (36). The scheme reduced social isolation and improved socialisation into the physical therapy career. The mentoring faculty reported increases in their cross cultural capital and professional growth (36). The study was robustly conducted but had a small sample size and impact on academic performance was not recorded.

#### Multilevel Interventions

King et al’s 2012 non-experimental cross-sectional study assessed whether diversity training reduces ethnic discrimination at individual and organisational levels through national surveys in the UK. The research highlighted that training method and content varies between NHS trusts, despite NHS guidance, and diversity training reduces the likelihood of discrimination and organisations can benefit from effective diversity programmes (50). However, the study methodology was poor with risks of bias.

Robinett et al conducted an observational study from 2017-2019 to mitigate bias in US medical school admissions. Interventions targeting the interview process and admissions team were assessed including: interviewer unconscious bias training, focused recruitment strategies and increased diversity on the admissions committee. There was an increase in underrepresented minority (URM) applicants accepted each year, in 2019 27.7% of accepted applicants identified as URM, compared to 49.3% in 2020 (51). The greatest impact was noted after they implemented a multilevel approach, as initially they only held unconscious bias training for the interviewers. Guh et al conducted a similar observational study looking at increasing racial diversity in a US residency programme. The interventions targeted the admissions team and included changes in interview scoring rubrics, targeted recruitment and a race and medicine workshop exploring bias and privilege. The study ran from 2014 to 2018 and resulted in a 19% increase in faculty and 40% increase in residents identifying as URMs (52). However, both papers are low quality, with high risks of bias.

Weech et al’s pre-post quasi-experimental study assessing the impact of a multifaceted cultural competency initiative on hospital performance metrics was the highest quality included paper. This was one of two studies with a control group. Four hospitals participated in the study over a 3.5 years period; two served as the control group and the others implemented the intervention. At an organisational level, focus was on improving diversity leadership, strategic human resource management, and patient cultural competency. At an individual level, the intervention targeted diversity attitudes, implicit bias, and racial/ethnic identity status. For the organisational and individual outcomes, hospitals with the interventions outperformed the control groups in both (37). The organisational outcome for increasing diversity in leadership was measured by Diversity Leadership and Cultural Competence Assessment and the Cultural Competency Assessment Tool for Hospitals (CCATH) and intervention hospitals experienced higher changes in scores. The individual outcome for reduction of implicit bias was measured by the Implicit Association Test (IAT) and found greater reduction in the intervention hospitals. Specific figures were not given for outcomes, limiting the quality of the paper as raw data are not available. The results show that a systems approach is effective at creating change in diversity and cultural competence practices in hospitals.

## Discussion

### Summary of main findings

This review of the international literature from 1970 to 2022, highlights an evidence gap for the effectiveness of antiracist interventions for healthcare staff, especially at an institutional level. Included studies were of poor quality, lacked long-term data and only four were from the UK. Most interventions focused on the personally mediated level (n=9) and were workshop based (n=8). All the personally mediated and multilevel interventions reported positive outcomes based on their interventions, however due to the poor methodological quality the results are not robust. Three studies assessed the effectiveness of institutional interventions with some indication that a diversity initiative could be successful though data were dated.

### Strengths and limitations

This is the first review examining antiracist interventions for healthcare staff in OECD countries. The protocol was published on PROSPERO, the papers were critically appraised using the most appropriate checklists and seven databases were systematically searched. The search was conducted following the Cochrane guidelines and in consultation with information scientists. Stakeholders supported the process to help interpret and validate findings.

Only 20% of the title and abstract screening was double reviewed, however this is in line with rapid review guidance (30). Citation searching was not extensive and there are risks of publication bias as no grey literature was included. The studies included were all low quality therefore knowledge about intervention effectiveness is inconclusive.

### Comparison with the literature

In 2021, NHS England published “Making antiracism a reality” where ‘diversity initiatives’ and ‘mentorship schemes’ were listed as interventions, however this review has shown that the evidence base behind them is poor (53). Multiple literature reviews have noted the inconsistencies between formal policies and their effective implementation (9, 21). In 1984, the Commission for Racial Equality (CRE) released guidelines about good practice for employers and in 1998, NHS trusts were surveyed about the implementation of their racial equality policies. Most had formal equal opportunity policies but only 5% implemented their action plans (9). The problems are ongoing and a survey by the Healthcare Commission in 2006 indicated a suspicion of “widespread non-compliance” across the NHS with regards to race relations legislation (54). This review highlights the need for further research to evaluate the impact of current policies and revise them as necessary.

The above “non-compliance” represents the wider issue for antiracism interventions - there is much information about potential interventions but few evaluated data. Various healthcare organisations, such as Kings Fund, a policy think tank, and the BMA, doctors union, publish reports about racial discrimination listing ‘successful’ interventions but use anecdotal evidence and include no methodology (55, 56).

The overall lack of institutional interventions is a key finding as research on racism often focuses on the interpersonal level with little emphasis on institutions (57). Racism manifests at different levels and ‘individual behaviour is shaped by organisational culture and practice’ (58), yet the interventions are disproportionately personally mediated.

Effective antiracist institutional interventions are critical as racism is based on power relationships between dominant and subordinate groups (59). Historical racism, such as colonialism, has shaped modern society and created racial inequities which has bled into our institutions. NHS Workforce Race Equality Standard (WRES) data showed white applicants were 1.46 times more likely to be appointed from shortlisting across all posts, while BAME staff were more likely to be formally disciplined by a factor of 1.22 (2).

The included papers lacked long-term follow-up, therefore it is unclear if the interventions have a continued impact and this requires evaluation. Lai et al examined whether implicit bias interventions had a lasting positive impact on American undergraduate students. While there were immediate positive outcomes the effects diminish over time (60). Furthermore, little is known about behavioural changes outside of the intervention setting (41, 44, 49, 50).

## Conclusion

There is a lack of robust evidence for the effectiveness of antiracist interventions targeted at healthcare staff, especially at an institutional level. Research is required to evaluate whether current antiracist policy and practice is effective, especially long-term. The lack of literature from the UK highlights a research gap. Discussions surrounding antiracism in healthcare need to move away from anecdotal evidence and towards high quality, evidence-based interventions which are adopted routinely.

## Data Availability

All data produced in the present study are available upon reasonable request to the authors

## Acknowledgements

The authors would like to acknowledge Rhian Jones and Katie Laugharne for their time, expertise and advice.

## Supplementary Material

### Search Strategy

| **Antiracism** | **Ethnic Minority** | **Healthcare Staff** |
| --- | --- | --- |
| (racism or race or racial) adj3<br>(discriminat* or bias* or prejudice*)<br>antiraci*<br>Anti raci*<br>Anti-raci* | Ethnic group*<br>Ethnic minorit*<br>Racial minorit*<br>Minorit* group*<br>Ethnicit*<br>BAME<br>Mulit-racial<br>Biracial<br>Bi-racial<br>(Mixed race)<br>(Black) ajd3 (African or Caribbean or Other)<br>(British) adj3 (Asian or African or Black)<br>Asian*<br>Indian*<br>Pakistan*<br>Bangladesh*<br>Chines*<br>Arab*<br>African*<br>Caribbean* | (healthcare or medical) adj3 (staff or worker* or workforce or faculty)<br>(health care) adj3 (staff or worker* or workforce or faculty)<br>(hospital) adj3 (staff or worker* or workforce or employee* or personnel)<br>Physician*<br>Health personnel* (use the subject heading)<br>NHS employee*<br>Hospital worker*<br>Healthcare worker*<br>Hospital staff<br>(NHS) adj3 (staff or employee*)<br>Physician*<br>Doctor*<br>Consultant*<br>Healthcare professional*<br>Nurse*<br>Paramedic*<br>Physiotherapist*<br>Occupational therapist*<br>Psychotherapist*<br>Physical therapist*<br>Radiologist*<br>Allied Health Personnel*<br>Caregiver*<br>Anaesthetist* |

### Supplementary material available on request

The data extraction tables are available on request to the corresponding author, Kismet Lalli.

## References

1. GOV.UK. Ethnic diversity of public sector workforces. 2019 [accessed 5.4.22]. Available from: https://www.ethnicity-facts-figures.service.gov.uk/summaries/public-sector-workforces#public-sector-staff-change-over-time

2. NHS. NHS workforce race and equality standard: 2019 data analysis report for NHS trusts. 2020 [accessed 5.4.22]. Available from: https://www.england.nhs.uk/wp-content/uploads/2020/01/wres-2019-data-report.pdf

3. Race Equality Foundation. The recruitment and retention of black and minority ethnic staff in the National Health Service. London 2007 [accessed 5.4.22]. Available from: http://raceequalityfoundation.org.uk/wp-content/uploads/2018/03/health-brief4.pdf

4. Doyin A, Kline R, Ochieng M. Fair to Refer?: GMC; 2019. Available from: https://www.gmc-uk.org/-/media/documents/fair-to-refer-report_pdf-79011677.pdf

5. Bhatia M. COVID-19 and BAME Group in the United Kingdom. The International Journal of Community and Social Development. 2020;2(2):269–72. doi: 10.1177/2516602620937878

6. Raleigh VS. Ethnic differences in covid-19 death rates. BMJ. 2022;376:o427. doi: 10.1136/bmj.o427

7. McKenzie K, Bhui K. Institutional racism in mental health care. BMJ. 2007;334(7595):649–50. doi: 10.1136/bmj.39163.395972.80

8. Race Equality Foundation. Experiences of bullying and racial harassment among minority ethnic staff in the NHS. 2009 [accessed 5.4.22]. Available from: https://raceequalityfoundation.org.uk/wp-content/uploads/2018/03/health-brief14.pdf

9. Kalra VS, Abel P, Esmail A, Kalra VS, Abel P, Esmail A. Developing leadership interventions for black and minority ethnic staff: A case study of the National Health Service (NHS) in the U.K. Journal of Health Organization & Management. 2009;23(1):103–18. doi: 10.1108/14777260910942588

10. Jones-Berry S. Is racism in the NHS being tackled effectively – or is it getting worse?: Trivialising racism as a ‘minority issue’ ignores its wider impact on staff, organisations and patients. Nursing Standard. 2019;34(10):19–21. doi: 10.7748/ns.34.10.19.s10

11. Kendi I. How to be an Antiracist. London, England: Bodley Head; 2019.

12. Calliste AM, Dei GJS. Power, knowledge and anti-racism education: A critical reader: Fernwood; 2000.

13. Hassen N, Lofters A, Michael S, Mall A, Pinto AD, Rackal J. Implementing Anti-Racism Interventions in Healthcare Settings: A Scoping Review. International Journal of Environmental Research and Public Health. 2021;18(6) doi: 10.3390/ijerph18062993

14. Brathwaite AC, Versailles D, Juudi-Hope D, Coppin M, Jefferies K, Bradley R, et al. Tackling discrimination and systemic racism in academic and workplace settings. NURSING INQUIRY. doi: 10.1111/nin.12485

15. Carter BM, Phillips BC. Revolutionizing the Nursing Curriculum. Creative Nursing. 2021;27(1):25–30. doi: 10.1891/CRNR-D-20-00072

16. Brown D. Providing an inclusive and safe environment. Practice Nursing. 2020;31(2):57–. doi: 10.12968/pnur.2020.31.2.57

17. Foster S. What it means to be an ally. British Journal of Nursing. 2021;30(7):453–. doi: 10.12968/bjon.2021.30.7.453

18. Jones-Berry S. BME representation: how to speed up the ‘glacial’ pace of change in the NHS. Nursing Standard. 2018;33(7):19–21. doi: 10.7748/ns.33.7.19.s11

19. Trueland J. How you can tackle casual racism and microaggression in the NHS: The steps every nurse can take to confront unconscious bias and be an authentic ally. Nursing Management - UK. 2020;27(5):14–7. doi: 10.7748/nm.27.5.14.s6

20. Watson L, Malcolm M. Racism as a Preventable Harm. Nursing Administration Quarterly. 2021;45(4):302–10. doi: 10.1097/NAQ.0000000000000495

21. Culley L. Equal opportunities policies and nursing employment within the British National Health Service. JOURNAL OF ADVANCED NURSING. 2001;33(1):130–7. doi: 10.1046/j.1365-2648.2001.01646.x

22. Joseph OR, Flint SW, Raymond-Williams R, Awadzi R, Johnson J. Understanding Healthcare Students’ Experiences of Racial Bias: A Narrative Review of the Role of Implicit Bias and Potential Interventions in Educational Settings. International journal of environmental research and public health. 2021;18(23) doi: 10.3390/ijerph182312771

23. Parsons S. Addressing Racial Biases in Medicine: A Review of the Literature, Critique, and Recommendations. INTERNATIONAL JOURNAL OF HEALTH SERVICES. 2020;50(4):371–86. doi: 10.1177/0020731420940961

24. Priest N, Esmail A, Kline R, Rao M, Coghill Y, Williams DR. Promoting equality for ethnic minority NHS staff—what works? BMJ: British Medical Journal. 2015;351(8019):16–8.

25. Tajeu GS, Halanych J, Juarez L, Stone J, Stepanikova I, Green A, et al. Exploring the association of healthcare worker race and occupation with implicit and explicit racial bias. Journal of the National Medical Association. 2018;110(5):464–72. doi: 10.1016/j.jnma.2017.12.001

26. Aggarwal NK, Cedeno KE, Lam P, Guarnaccia P, Lewis-Fernandez R. PERCEPTIONS OF IMPLEMENTING CULTURALLY COMPETENT COMMUNICATION AMONG PATIENTS, CLINICIANS, AND ADMINISTRATORS: A MIXED-METHODS EXPLORATORY STUDY. Journal of Cultural Diversity. 2018;25(1):3–11.

27. Valdez P. What can I do? A call for pediatric providers to engage in antiracism and social justice for the health of their patients. Journal of Developmental and Behavioral Pediatrics. 2020;41(7):504–5. doi: 10.1097/DBP.0000000000000853

28. Page MJ MJ, Bossuyt PM, Boutron I, Hoffmann TC, Mulrow CD. The PRISMA 2020 statement: an updated guideline for reporting systematic reviews. The BMJ. 2021;372(71) doi: 10.1136/bmj.n71

29. Cochrane Handbook for Systematic Reviews of Interventions version 6.3. In: Higgins JPT TJ, Chandler J, Cumpston M, Li T, Page MJ, Welch VA, editor.: Cochrane; 2022.

30. Garritty C GG, Kamel C, King VJ, Nussbaumer-Streit B,, Stevens A HC, Affengruber L. Cochrane Rapid Reviews. Interim Guidance from the Cochrane Rapid Reviews Methods Group. Reviews CR; 2020. Available from: https://methods.cochrane.org/rapidreviews/sites/methods.cochrane.org.rapidreviews/files/public/uploads/cochrane_rr_-_guidance-23mar2020-final.pdf

31. Leatherdale ST. Natural experiment methodology for research: a review of how different methods can support real-world research. International Journal of Social Research Methodology. 2019;22(1):19–35. doi: 10.1080/13645579.2018.1488449

32. National Heart Lungs and Blood Institute. Study Quality Assessment Tools. 2021 [accessed 22.03.22]. Available from: https://www.nhlbi.nih.gov/health-topics/study-quality-assessment-tools

33. Joanna Briggs Institute. Checklist for Qualitative Research. 2020 [accessed 25.3.22]. Available from: https://jbi.global/critical-appraisal-tools

34. Joanna Briggs Institute. Checklist for Quasi Experimental Studies. 2020 [accessed 25.3.33]. Available from: https://jbi.global/critical-appraisal-tools

35. Hong QN PP, Fàbregues S, Bartlett G, Boardman F, Cargo M, Dagenais P, Gagnon, M-P GF, Nicolau B, O’Cathain A, Rousseau M-C, Vedel I. Mixed Methods Appraisal Tool (MMAT). Canada: 2018 [accessed 25.3.33]. Available from: http://mixedmethodsappraisaltoolpublic.pbworks.com/w/file/fetch/127916259/MMAT_2018_criteria-manual_2018-08-01_ENG.pdf

36. Naidoo K. Networked mentoring to promote social belonging among minority doctor of physical therapy students. Dissertation Abstracts International: Section B: The Sciences and Engineering. 2021;82(10-B):No-Specified.

37. Weech-Maldonado R, Dreachslin JL, Patien Epané J, Gail J, Gupta S, Wainio JA. Hospital cultural competency as a systematic organizational intervention: Key findings from the national center for healthcare leadership diversity demonstration project. Health Care Management Review. 2018;43(1):30–41. doi: 10.1097/HMR.0000000000000128

38. Wusu MH, Tepperberg S, Weinberg JM, Saper RB. Matching Our Mission: A Strategic Plan to Create a Diverse Family Medicine Residency. Family Medicine. 2019;51(1):31–6. doi: 10.22454/FamMed.2019.955445

39. Bagilhole B, Stephens M. Management Responses to Equal Opportunities for Ethnic Minority Women Within an NHS Hospital Trust. Journal of Social Policy. 1999;28(2):235–48. doi: 10.1017/S0047279499005541

40. Johns N. Ethnic Diversity Policy: Perceptions within the NHS. Social Policy & Administration. 2004;38(1):73–88. doi: 10.1111/j.1467-9515.2004.00377.x

41. Bheenuck S, Miers M, Pollard K, Young P. Race equality education: implications of an audit of student learning. Nurse Education Today. 2007;27(5):396–405. doi: 10.1016/j.nedt.2006.06.003

42. Wilson-Mitchell K, Handa M. Infusing Diversity and Equity Into Clinical Teaching: Training the Trainers. Journal of Midwifery & Women’s Health. 2016;61(6):726–36. doi: 10.1111/jmwh.12548

43. Sotto-Santiago S, Mac J, Duncan F, Smith J. “I Didn’t Know What to Say”: Responding to Racism, Discrimination, and Microaggressions With the OWTFD Approach. MedEdPORTAL: the journal of teaching and learning resources. 2020;16:10971. doi: 10.15766/mep_2374-8265.10971

44. Stone J, Moskowitz GB, Zestcott CA, Wolsiefer KJ. Testing Active Learning Workshops for Reducing Implicit Stereotyping of Hispanics by Majority and Minority Group Medical Students. STIGMA AND HEALTH. 2020;5(1):94–103. doi: 10.1037/sah0000179

45. White-Davis T, Edgoose J, Speights JSB, Fraser K, Ring JM, Guh J, et al. Addressing Racism in Medical Education An Interactive Training Module. Family Medicine. 2018;50(5):364–8. doi: 10.22454/FamMed.2018.875510

46. Steed R. Attitudes and beliefs of occupational therapists participating in a cultural competency workshop. Occupational Therapy International. 2010;17(3):142–51. doi: 10.1002/oti.299

47. Dancis J, Coleman BR. Transformative dissonant encounters: Opportunities for cultivating antiracism in White nursing students. Nursing Inquiry. 2021 doi: 10.1111/nin.12447

48. Rezaei AR. Validity and reliability of the IAT: Measuring gender and ethnic stereotypes. Computers in Human Behavior. 2011;27(5):1937–41. doi: 10.1016/j.chb.2011.04.018

49. Roswell RO, Cogburn CD, Tocco J, Martinez J, Bangeranye C, Bailenson JN, et al. Cultivating Empathy Through Virtual Reality: Advancing Conversations About Racism, Inequity, and Climate in Medicine. ACADEMIC MEDICINE. 2020;95(12):1882–6. doi: 10.1097/ACM.0000000000003615

50. King EB, Dawson JF, Kravitz DA, Gulick LMV. A multilevel study of the relationships between diversity training, ethnic discrimination and satisfaction in organizations. Journal of Organizational Behavior. 2012;33(1):5–20. doi: 10.1002/job.728

51. Robinett K, Kareem R, Reavis K, Quezada S. A multi-pronged, antiracist approach to optimize equity in medical school admissions. Medical education. 2021;55(12):1376–82. doi: 10.1111/medu.14589

52. Guh J, Harris CR, Martinez P, Chen FM, Gianutsos LP. Antiracism in Residency: A Multimethod Intervention to Increase Racial Diversity in a Community-Based Residency Program. Family medicine. 2019;51(1):37–40. doi: 10.22454/FamMed.2019.987621

53. NHS England and NHS Improvement. Making anti-racism a reality. 2021 [accessed 4.4.22]. Available from: https://www.england.nhs.uk/east-of-england/wp-content/uploads/sites/47/2021/09/NHS-England-and-NHS-Improvement-East-of-England-antiracism-strategy.pdf

54. Healthcare Commission. Healthcare Commission Annual Report 2006/2007. 2006 [accessed 5.4.22]. Available from: https://assets.publishing.service.gov.uk/government/uploads/system/uploads/attachment_data/file/250569/0913.pdf

55. The Kings Fund. Workforce race inequalities and inclusion in NHS providers. England, 2020 [accessed 5.4.22]. Available from: https://www.kingsfund.org.uk/sites/default/files/2020-07/workforce-race-inequalities-inclusion-nhs-providers-july2020.pdf

56. British Medical Association. A charter for medical schools to prevent and address racial harassment. 2020 [accessed 5.4.22]. Available from: https://www.bma.org.uk/media/2030/bma-med-school-charter-implementation.pdf

57. Bailey ZD, Krieger N, Agénor M, Graves J, Linos N, Bassett MT. Structural racism and health inequities in the USA: evidence and interventions. The Lancet. 2017;389(10077):1453–63. doi: 10.1016/S0140-6736(17)30569-X

58. Griffith DM, Mason M, Yonas M, Eng E, Jeffries V, Plihcik S, et al. Dismantling institutional racism: theory and action. Am J Community Psychol. 2007;39(3-4):381–92. doi: 10.1007/s10464-007-9117-0

59. Blalock HM. A Power Analysis of Racial Discrimination. Social Forces. 1960;39(1):53–9. doi: 10.2307/2573575

60. Lai CK, Skinner AL, Cooley E, Murrar S, Brauer M, Devos T, et al. Reducing implicit racial preferences: II. Intervention effectiveness across time. J Exp Psychol Gen. 2016;145(8):1001–16. doi: 10.1037/xge0000179

